# Oxygenator use in Ventricular Assist Devices from the Advanced Cardiac Therapies Improving Outcomes Network (ACTION) Registry

**DOI:** 10.1101/2025.06.10.25329341

**Authors:** Ahmed S. Said, Mary Mehegan, Anna Joong, Adam Morrison, David Sutcliffe, Joseph Philip, Muhammad Shezad, Rachel E Harris, Matthew Amidon, Christopher Knoll, Bethany Wisotzkey, Mohammed Absi, Megan Wilde, Arene Butto, John Dykes, Kae Watanabe, Matthew Zinn, Lindsay May, Christina VanderPluym, Elizabeth Braunlin, Chesney Castleberry, Shriprasad Deshpande, Angela Lorts, Lydia K Wright, Matthew O’Connor, Deepa Mokshagundam, Edon J. Rabinowitz

## Abstract

**Objective:** To describe the utilization patterns and outcomes of pediatric patients supported with oxygenators during their ventricular assist device (VAD) course.

**Design:** Multi-center, retrospective cohort study using data from the Advanced Cardiac Therapies Improving Outcomes Network (ACTION) registry.

**Setting:** Data query from the ACTION registry, which collects clinical data on pediatric patients with end-stage heart disease, reported cases of combined oxygenator-VAD use.

**Patients:** Sixty-one pediatric patients (27 female) from 21 contributing ACTION centers were supported on oxygenators during their VAD course between 2013-2024. The median age was 272 days (IQR 68-1334), and the median weight was 7.9kg (IQR 4.4-12). The majority had congenital heart disease (CHD), with 66% having single-ventricle pathology.

**Interventions:** None.

**Measurement and Main Results:** The study population demonstrated high illness severity at the time of VAD implant, with 97% requiring inotropic support, 79% requiring mechanical ventilation, and 46% on ECMO. The total median duration of VAD support was 45 days (IQR 16-163). Of the 61 patients, 44% achieved transplant or recovery. The mortality rate was high at 52%, and was associated with younger age, smaller size, CHD and pre-implant use of neuromuscular blocking agents. Adverse events included infection (36%), major bleeding (36%), central nervous system injuries (21%), and dialysis (20%). None demonstrated clinically significant association with oxygenator support timing, but mortality was associated with higher adverse events rates, particularly pulmonary hemorrhage, dialysis, and infections (especially mediastinitis).

**Conclusion:** Pediatric patients requiring oxygenators during VAD therapy exhibit high illness severity and frequent adverse events. Mortality is high and associated with younger age, smaller size, CHD, pre-implant use of neuromuscular blocking agents as well frequency of on-device adverse events, particularly pulmonary hemorrhage, dialysis, and infections. Oxygenator support likely reflects disease severity rather than directly contributing to mortality. Further investigation is needed to optimize patient selection and management strategies.

**Research In Context:**

- Over the past two decades, pediatric VAD utilization has increased. Support from collaborative networks like ACTION has enabled investigation of nuanced strategies, such as temporary oxygenator use, providing a platform to evaluate feasibility, safety, and patient-specific considerations across diverse populations.
- In adult populations, oxygenator use alongside VADs has been employed for respiratory failure, ARDS, RV support, and as a bridge to lung transplant or transition from ECMO. However, these strategies have not been systematically studied in pediatric cohorts until now.
- This study presents the most comprehensive review of pediatric (or adult) oxygenator-VAD support to date, characterizing patient populations, utilization patterns, and associated outcomes. It provides foundational data for guiding future research into management strategies in this high-risk group.

**At the Bedside:**

- Pediatric patients requiring oxygenator support during VAD therapy were typically younger, smaller, and had complex congenital heart disease, particularly single-ventricle physiology. These characteristics were associated with higher mortality and reflect a population with profound critical illness at baseline.
- This cohort experienced high rates of adverse events, including major bleeding, infections, dialysis, and neurologic injury. While oxygenator timing was not independently associated with outcomes, mortality exceeded 50% and was linked to complications such as dialysis, pulmonary hemorrhage, and infections.
- Clinicians should view oxygenator-VAD support as a surrogate marker for extreme illness severity rather than a direct contributor to poor outcome. This study highlights the need for tailored management strategies and informs clinician and family decision-making in this high-risk population.

## INTRODUCTION

The utilization of ventricular assist devices (VADs) in pediatric populations has significantly expanded over the past two decades, driven in part by the establishment of large, inclusive collaborative networks such as the Advanced Cardiac Therapies Improving Outcomes Network (ACTION)^1,2^. These multicenter collaborations have facilitated a deeper understanding of nuanced approaches to VAD use in children. One notable strategy is the integration of oxygenators with durable VADs.

Temporary oxygenator use alongside VAD support has been well-documented in adults under various clinical circumstances. These include managing respiratory failure^3-5^, cardiogenic shock^3,6,7^, as part of underlying right ventricular (RV) support strategies^8-10^; serving as a bridge to lung transplantation^11,12^; addressing COVID-19-related cardiopulmonary complications/acute respiratory distress syndrome (ARDS)^13,14^; and transitioning from peripheral extracorporeal membrane oxygenation (ECMO) to durable VAD support^15^.

In pediatrics, however, descriptions of oxygenator incorporation into VAD strategies are limited to single-center case reports or isolated mentions within broader case series. Despite this, the feasibility and benefits of this approach have been highlighted, including avoiding prolonged toxic ventilator support, additional surgical procedures for separate ECMO cannulation, increased thrombotic risk from vessel manipulation, and the complexities of managing dual mechanical circulatory support (MCS) devices^16^. Pediatric cases reporting effective oxygenator use with VADs have involved respiratory failure due to infection^17^, ARDS^18,19^, and multifactorial etiologies^20-24^. These cases have typically been described as bridges to pulmonary recovery or as part of paracorporeal artificial lung support bridging to lung transplantation^25-28^.

These reports contribute to our understanding of VAD strategies, care, and outcomes, however the limited data available suggest variations in care practices and the timing of oxygenator deployment between centers. To address this knowledge gap, we analyzed data from the ACTION registry^1^. Our objectives were to describe pre-implant clinical characteristics, VAD implantation strategies, oxygenator utilization patterns, and survival outcomes at transplant.

## MATERIALS & METHODS

We conducted a retrospective analysis of the ACTION registry, including all pediatric patients supported with VADs between June 2013 and June 2024. Patients without documented oxygenator use or insufficient data were excluded. This study was exempt from review by the Washington University in St. Louis Institutional Review Board (IRB) because the analyzed data were fully anonymized, and no authors had access to identifiable data. The ACTION registry itself is approved by the IRB at Cincinnati Children’s Hospital, where the data are maintained.

Data collection encompassed clinical and surgical characteristics before and after VAD and oxygenator support, as well as adverse events, which were defined according to the established ACTION registry criteria^1^. Data abstraction was conducted by trained registry personnel and subsequently adjudicated for selected adverse events by two blinded reviewers and a committee to ensure consistency with registry definitions. Detailed definitions of all study variables have recently been published^29^.

Statistical analyses were performed using GraphPad version 9 (GraphPad Software, CA). Descriptive statistics, including frequencies and percentages for categorical variables and medians with interquartile ranges (IQRs) for continuous variables, were used to summarize patient characteristics. Predefined variables were analyzed both individually and as composite measures, with comparisons made between early oxygenator users (defined as oxygenator initiation within the immediate post-implant period; <24 hours) and late oxygenator users (defined as oxygenator initiation >24 hours after VAD implantation) as well as survivors and non-survivors. Group comparisons of quantitative variables were conducted using the Mann-Whitney test, while categorical variables were analyzed using the Chi-square test. A p-value of <0.05 was considered statistically significant.

## RESULTS

### Pre-implant patient characteristics and support

At the time of this study, the ACTION registry included data from 45 centers, with 61 patients (27 female) from 21 centers meeting the inclusion criteria between 2013 and 2024 (**Table 1 & digital supplement to table 1**). The median age at VAD implantation was 272 days [IQR 68–1334], with a median weight of 7.9 kg [IQR 4.4–12] and a body surface area (BSA) of 0.39 m^2^ [IQR 0.26– 0.54].

The majority of patients (79%) had underlying congenital heart disease (CHD), most commonly single ventricle heart disease (66%). The remaining 21% were predominantly cases of cardiomyopathy or myocarditis. Consequently, 79% of patients had undergone at least one prior sternotomy, with 52% having had two or more prior sternotomies.

The Interagency Registry for Mechanically Assisted Circulatory Support (INTERMACS) profiles at the time of VAD implantation were distributed as follows: Profile 1 in 49%, Profile 2 in 48%, and Profile 3 in 3%. A history of cerebrovascular accidents (CVA) prior to VAD implantation was reported in 16% of patients, while an additional 15% had experienced non-CVA neurological injuries, such as intracranial bleeding, before VAD placement.

In the 7 days preceding VAD implantation, patients required substantial medical support. Nearly half (46%) were transitioned directly from ECMO. The majority (97%) were receiving inotropic support, with 56% on two or more inotropes at the time of implantation. Mechanical ventilation was utilized in 79% of patients, and neuromuscular blockade was employed in 43%. Additionally, most patients (72%) were dependent on total parenteral nutrition (TPN), while 8% required dialysis during this period.

ECMO use immediately preceding VAD implantation, was associated with early oxygenator use post-implantation (*p=*0.0010). Pre-implant characteristics associated with post-implant mortality included younger age (median 201 vs 879 days, *p=*0.0143), lesser weight (median weight 5.6kg vs 10.5kg, *p=*0.0156), and smaller BSA (0.30m^2^ vs 0.51m^2^, *p=*0.0114). Post-implant mortality was also more likely to be seen in patients with CHD (*p=*0.0466) and those receiving neuromuscular blocking agents in the week leading up to VAD implantation (*p=*0.0402).

### VAD implantation strategies

Device utilization strategies varied across centers. Most patients (56%) underwent VAD implantation as a bridge to transplantation, while a notable proportion were implanted as a bridge to candidacy (26%), recovery (16%), or as destination therapy (2%).

Initial device selection was nearly evenly split between paracorporeal continuous flow devices (51%) and paracorporeal pulsatile flow devices (49%). All pulsatile devices were transitioned to continuous flow devices likely to accommodate oxygenator use.

Concomitant surgical procedures were performed in 44% of cases during VAD implantation. Of note, early oxygenators were employed in 34% of patients (21 cases), likely as part of an acute postoperative recovery strategy. For the remaining 66% of patients (40 cases), late oxygenators were utilized.

### VAD course & outcomes

The study cohort remained on VAD support for a median duration of 45 days [IQR 16–163] Overall, survival to transplant while on support was poor, achieved in only 27 of 61 patients (44%) (**Figure 1**). All patients who transplanted or recovered were able to be separated from the oxygenator and remain on VAD support alone through heart transplantation. Mortality on device was nearly identical between early and late oxygenator users, with 52% and 53% of patients, respectively, dying while on support. Of the 32 patients who died on device, causes included, multi-organ dysfunction (n=8, 25%), overwhelming infections (n=5, 16%), CVA (n=2, 6%), renal failure changing organ candidacy and leading to withdrawal of life-sustaining support (n=2, 6%), pulmonary venous stenosis (n=1, 3%) major bleeding (n=1, 3%). Unknown causes of death (n=13, 41%) were seen at a high rate, which may be reflective of an ACTION registry limitation in which registry data is limited after patients are transitioned to ECMO (i.e. supported with an oxygenator).

**Figure 1:**
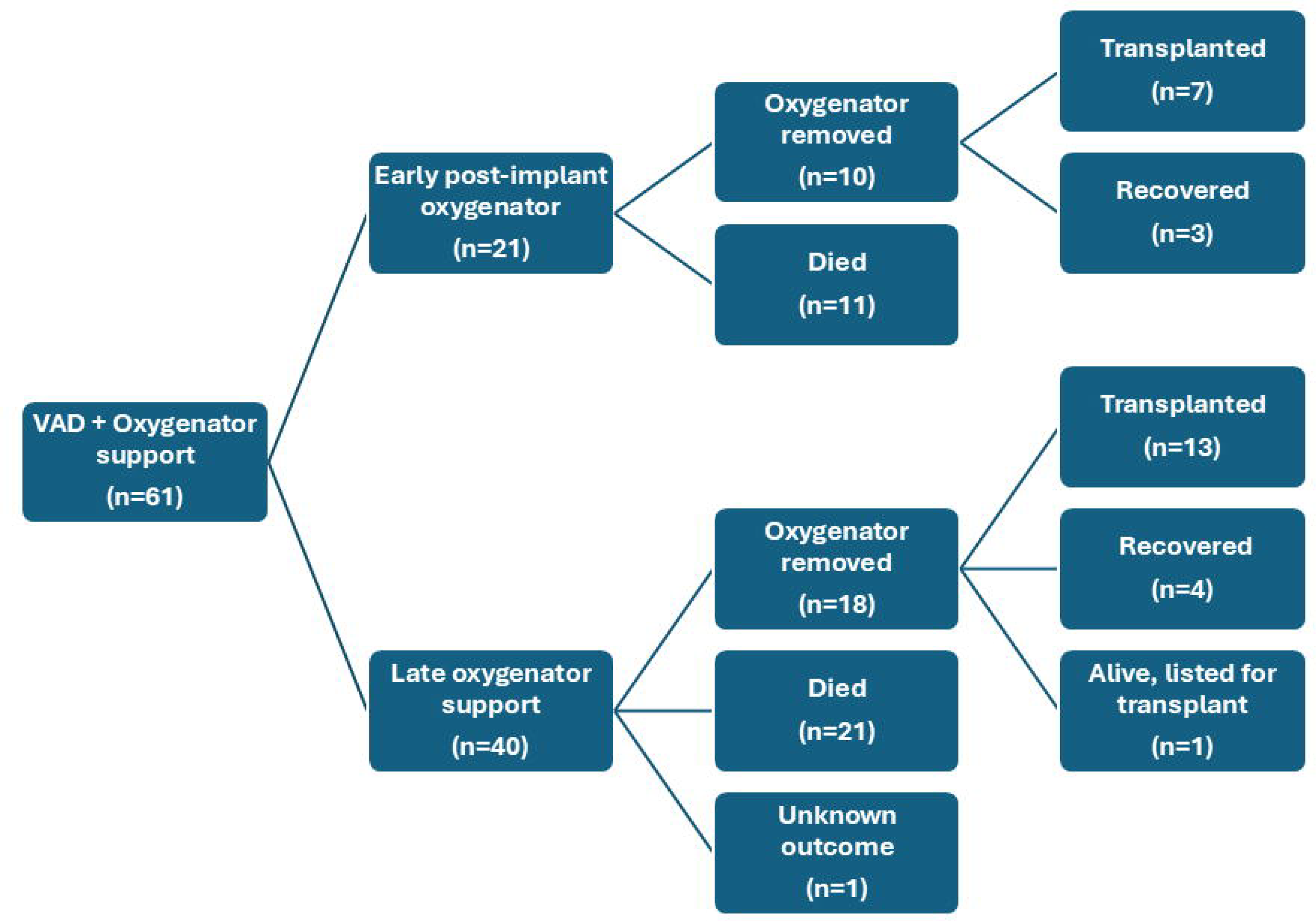
Among the 61 patients identified in the ACTION registry, 21 utilized oxygenators in the immediate post-VAD implant period (<24 hours post-implant). Of these, 11 patients (52%) died, 7 (33%) had their oxygenators successfully removed and subsequently achieved transplantation, and 3 (13%) had recovered and explanted. Late oxygenator support (>24 hours post-implant) was reported in 40 patients. Within this group, 21 patients (53%) died, 13 (33%) were able to transition back to VAD support alone without the oxygenator and ultimately underwent successful transplantation, and 4 (10%) had recovered. Unknown outcomes were reported in 1 patient (3%), likely reflecting limitations in registry reporting. At the time of this analysis, 1 patient is alive and listed for transplant (3%).

A total of 157 adverse events were reported in 51 patients (84%). These events are summarized in **Table 2**. Central nervous system (CNS) injuries were reported in 13 patients (21%), with a total of 15 CNS events, including ischemic CVA (n=6), hemorrhagic CVA (n=2), and intracranial hemorrhage (n=7). CNS events occurred at a median of 14.5 days post-implant [IQR 2.5–22.5]. Other adverse events included 23 major bleeds in 22 patients (36%), occurring at a median of 27.5 days post-implant [IQR 3–51], all of which required transfusions, and 7 of which underwent surgical intervention to control hemorrhaging. Dialysis was required in 12 patients (20%). A total of 39 major infections were observed in 22 patients (36%), with bloodstream infections constituting nearly half of the reported events. Infections occurred more frequently in early oxygenator users (57%) compared to late oxygenator users (25%) (*p=*0.0130) and may be reflective of longer device days seen in early vs late oxygenator users. Hemolysis was reported in 9 patients (15%) and occurred more commonly in early oxygenator users (33%) compared to late oxygenator users (5%) (*p=*0.0057), also perhaps reflective of longer device days seen in early vs late oxygenator users.

For most adverse events, no statistically significant differences were observed based on the timing of oxygenator use. However, multiple important factors were associated with those who died compared to those who achieved transplant or recovery. Each patient who died experienced adverse events during their VAD course compared to 70% of those who achieved transplantation or recovery (*p=*0.0001). Most notably, mortality was associated with post-implant pulmonary hemorrhage (*p=*0.0318), dialysis (*p=*0.0234), and infection (*p=*0.0062), particularly mediastinitis (*p=*0.0318).

## DISCUSSION

To our knowledge, this report represents the largest analysis of combined oxygenator-VAD support in pediatric (or adult) patients and is the first multi-center study to focus on this unique population. It provides valuable insights into current practice trends and VAD experiences for this particularly challenging group of patients. The key findings indicate that pediatric patients requiring oxygenators during VAD therapy exhibited high illness severity both before and after VAD implantation, with frequent adverse events and a high mortality rate of 52%. Complications and mortality appeared to occur irrespective of whether oxygenators were initiated <24 hours post-implant or later. Mortality, however, similar to traditional pediatric (non-oxygenator) VAD risk factors^30,31^, was more likely to be seen in those younger and smaller patients, with CHD and on neuromuscular blocking agents pre-implant. The frequency of on-device adverse events was associated with mortality, particularly pulmonary hemorrhage, dialysis, and infections (especially mediastinitis).

The higher rates of adverse events observed in early oxygenator users, such as infections and hemolysis, may be more attributable to the longer duration of device support in this group (median of 113 days compared to 34.5 days for late oxygenator users) rather than an inherently greater risk associated with early oxygenator use. Similarly, while oxygenator use itself does not appear to directly contribute to mortality, its use likely signifies the severity of the underlying illness and a generally poor prognosis.

Case series from adult VAD cohorts have previously described the use of oxygenators in conjunction with VADs, often referred to as “Oxy-VAD.” Some of these studies focused on short-term support, with a median duration of 8.5 days, aimed not at bridging to transplantation but as a strategy for respiratory recovery while simultaneously providing cardiac support^3^. This short-term approach has been described as lifesaving during pulmonary decompensation secondary to left heart failure, with the oxygenator serving to support gas exchange until conventional mechanical ventilation can be relied upon^4,5^. Additionally, some adult reports have suggested a hematologic advantage in combining oxygenators with VADs, noting a decrease in transfusion requirements^6^. However, this finding has not been consistently reported, with other studies indicating higher rates of bleeding complications, along with greater short-term complications such as prolonged intubation and referral for tracheostomy^7^. Similar to our findings, we suspect that these complications may reflect the underlying disease burden of the patient population and the decision to resort to oxygenators as a therapeutic strategy, rather than the oxygenator itself being inherently dangerous.

Other adult reports have focused on the use of oxygenators for temporary right-sided VAD support in conjunction with LVADs. This approach is typically used in the perioperative period, where right heart VAD support is completely removed along with the oxygenators when right heart recovery is observed^8,10^. Some studies have suggested a short-term survival benefit with this strategy^9^. However, long-term outcome differences between patients with or without the use of an oxygenator have not been clearly established. Short-term use of oxygenators in combination with VAD support have more recently been described during the COVID-19 pandemic, where systemic inflammation and ARDS were the hallmark of pulmonary complications^13,14^.

Strategies to transition peripheral ECMO support to more durable VADs, with temporary reliance on oxygenators during the transition period, have also been described^15^. These strategies are similar to those employed by the early oxygenator users in our study, where 76% were on pre-implant ECMO support. Intuitively, this approach may allow for stabilization during the transition phase, providing the necessary respiratory and circulatory support as patients are moved to chronic VAD therapy. However, in our study, high mortality rates, similar to late oxygenator users, were seen in early oxygenator users.

Adult medicine has also reported the use of “Oxy-VAD” as a long-term bridge to lung transplantation^11,12^, a strategy that constitutes most of the reports in the pediatric population. As an example, at Washington University in St. Louis, patients have been supported prior to lung transplantation with paracorporeal lung assist devices specifically to aid gas exchange in patients with severe interstitial lung disease and pulmonary hypertension^25-28^. Some of these patients have been supported without a mechanical ventilator, allowing for rehabilitation over extended periods, with some cases lasting up to 74 days. Importantly, this approach utilizes a pumpless system where blood flow through the oxygenator is driven by pressure gradients, with a pulmonary artery-to-left atrial configuration, instead of relying on a pump to assist in circulation. This experience is not represented in the ACTION registry.

Previous pediatric reports, likely represented in the ACTION registry, have described cases of respiratory failure due to infection^17^, ARDS^18,19^, and multifactorial etiologies^20-24^. The potential benefits of oxygenator use in VAD circuit include avoiding prolonged use of toxic ventilator settings, additional surgical procedures for separate ECMO cannulation, increased thrombotic risks from vessel manipulation, and the complexities of managing dual MCS devices^16^. Specific patient case reports have highlighted the incorporation of oxygenators into left heart VAD support in a patient with pneumonia and ARDS^17^. In this case, mechanical ventilatory support was so harmful that ventricular-ventricular interactions were impairing left heart filling, which in turn affected VAD filling and cardiac output. Oxygenator use was credited with easing ventilatory requirements, reducing right heart afterload, improving VAD filling, and lessening the need for vasoactive support. Other pediatric reports have described the use of oxygenator rescue therapy for acute pneumonia and ARDS in patients with right-sided VADs, forming part of an overall biventricular assist device (BiVAD) strategy for cardiac support^18,19^.

As reflected in this study and others^21,32^, oxygenator use is typically accomplished with continuous flow devices. However, there have been reports from Europe describing the incorporation of oxygenators into the outflow cannula of Berlin Heart EXCOR assist devices (Berlin Heart, Berlin, Germany)^20^. These reports, akin to our study, primarily involve patients with congenital heart disease, particularly those with single ventricle heart disease^21,23,24,32^. While not originally designed to evaluate oxygenator efficacy, these studies, like our analysis, do not conclusively demonstrate a survival benefit associated with oxygenator use. They do, however, underscore the predilection for adoption of oxygenators in single ventricle heart disease, a patient population predisposed to chronic hypoxia and impaired end-organ oxygen delivery during states of low cardiac output especially in the presence of concurrent pulmonary pathology. This single ventricle cohort represents a notably high-risk subset within VAD cohorts, irrespective of oxygenator utilization ^24,33^ with recent ACTION analyses reporting transplant rates ranging from 63% to 76%.

Adverse events were reported in the majority of patients, with complication rates higher than those previously reported for biventricular disease and slightly worse than those seen with single ventricle VAD support^24,33-35^. This is likely reflective of the particularly high-risk cohort described in the analysis, rather than an effect of the oxygenator itself. The occurrence of CVAs remained high in this study cohort, with rates similar to that seen in previous reports of single ventricle VAD support^24,33^, likely due to significant overlap in the patient populations. Chronic hypoxemia may be directly related to the high stroke rates, as it can lead to inappropriate vasoconstriction and vascular stasis at both the micro- and macro-vascular levels, combined with endothelial dysfunction and red blood cell deformation^36-40^. This chronically hypoxemic cohort might benefit from a more aggressive and individualized anti-thrombotic strategy to mitigate this risk, though this approach would need to be balanced against the frequency of major bleeding events, which were seen in over one-third of patients in this study.

This study was limited by its heterogeneous and modest sample size, as well as the quality of registry data. Despite the ACTION adjudication process, it is possible that not all relevant events related to patient outcomes were either entered by participating centers or captured by the current data fields. Specifically, a significant proportion of patients were reported to have an unknown cause of death, which may reflect the cessation of data entry after oxygenator use was initiated. For the same reasons, we suspect adverse events may be under-reported. Additionally, detailed data regarding the timing of intensive therapy cessation, such as the length of mechanical ventilation, was unavailable for analysis as a surrogate of effective VAD and/or oxygenator support. The study was descriptive in nature, lacking a comparison control group, and was not powered for meaningful univariable or multivariable analyses. The associations observed between early vs. late oxygenator users in the study may not represent strong signals in this small sample size, and further investigation is required. These limitations highlight the need for additional research to identify candidates who would most benefit from oxygenator incorporation during VAD therapy, as well as to determine the optimal timing for such support.

### Conclusion

In summary, our findings indicate that pediatric patients requiring oxygenators during VAD therapy experience high illness severity, frequent adverse events, and a high mortality rate (52%). Mortality and complications occurred regardless of when the oxygenator was initiated. Mortality rates were higher in younger and smaller patients as well as in those experiencing higher adverse event rates, particularly pulmonary hemorrhage, severe renal failure requiring dialysis, and infections, especially mediastinitis. Oxygenator use likely signals disease severity rather than independently influencing outcomes. Individualized consideration and further multi-centered investigation are required, as current best practice standards and consensus regarding oxygenator deployment and strategy are lacking.

## Data Availability

All data produced in the present study are available upon reasonable request to the authors with approval from the Advanced Cardiac Therapies Improving Outcomes Network (ACTION)

