## Supplementary material for "Oxygenator use in Ventricular Assist Devices from the Advanced Cardiac Therapies Improving Outcomes Network (ACTION) Registry": Table 1

**Digital Supplement Table 1: patient characteristics & pre-implant support**

| Variable<br>Median [IQR] | Total pts<br>n = 61 | In-Hospital Outcome | | P value | In-Hospital Outcome** | | P value |
|---|---|---|---|---|---|---|---|
| | | Early Oxygenator<br>n = 21 | Late Oxygenator<br>n = 40 | | Died<br>n=32 | Transplanted or Recovered<br>n=27 | |
| Age at implant, days | 272 [68-1334] | 154 [88-879] | 274 [65-1624] | 0.6341 | 201 [65-518] | 879 [149-2384] | ***0.0143** |
| Wt at implant, kg | 7.9 [4.4-12] | 6.8 [4-11.2] | 8 [4.4-15.7] | 0.4974 | 5.6 [4.2-9.8] | 10.5 [6.4-19.1] | ***0.0156** |
| BSA, m2 | 0.39 [0.26-0.54] | 0.31 [0.26-0.51] | 0.39 [0.26-0.67] | 0.3713 | 0.30 [0.25-0.45] | 0.51 [0.29-0.77] | ***0.0114** |
| Female, n (%) | 27 (44%) | 11 (52%) | 16 (40%) | 0.3550 | 14 (43%) | 13 (48%) | 0.7968 |
| Congenital Heart Disease, n (%) | 48 (79%) | 17 (81%) | 31 (78%) | 0.6339 | 29 (90%) | 19 (70%) | ***0.0466** |
| Single Ventricle Disease, n (%) | 40 (66%) | 14 (67%) | 26 (65%) | 0.8964 | 24 (75%) | 16 (59%) | 0.3677 |
| Single RV | 29 (48%) | 12 (57%) | 17 (42%) | 0.1696 | 18 (56%) | 11 (40%) | 0.2993 |
| Single LV | 11 (18%) | 2 (10%) | 9 (22%) | 0.1696 | 6 (19%) | 5 (18%) | 0.9999 |
| Non-congenital diagnosis | 13 (21%) | 3 (14%) | 10 (25%) | 0.4005 | 4 (12%) | 9 (33%) | 0.0544 |
| DCM / Myocarditis | 8 (13%) | 2 (10%) | 6 (15%) | - | 3 (9%) | 5 (19%) | - |
| RCM | 2 (3%) | 0 (0%) | 2 (5%) | - | 0 (0%) | 2 (7%) | - |
| HCM | 1 (2%) | 0 (0%) | 1 (3%) | - | 1 (3%) | 0 (0%) | - |
| LVNC | 1 (2%) | 1 (5%) | 0 (0%) | - | 0 (0%) | 1 (4%) | - |
| Transplant graft rejection | 1 (2%) | 0 (0%) | 1 (3%) | - | 1 (3%) | 0 (0%) | - |
| Prior Sternotomies, n (%) | 43 (70%) | 18 (86%) | 25 (62%) | 0.0589 | 26 (81%) | 17 (63%) | 0.0697 |
| 1 | 16 (26%) | 7 (33%) | 9 (41%) | - | 12 (38%) | 4 (15%) | - |
| 2 | 8 (13%) | 3 (14%) | 5 (13%) | - | 4 (13%) | 4 (15%) | - |
| 3 | 11 (18%) | 5 (24%) | 6 (15%) | - | 6 (19%) | 5 (18%) | - |
| ≥4 | 8 (13%) | 3 (14%) | 5 (13%) | - | 4 (13%) | 4 (15%) | - |
| INTERMACS profile at implant, n (%) | | | | | | | |
| 1 | 30 (49%) | 8 (38%) | 22 (55%) | 0.2831 | 19 (59%) | 11 (41%) | 0.1538 |
| 2 | 29 (48%) | 10 (48%) | 19 (48%) | 0.6020 | 12 (38%) | 17 (63%) | 0.0513 |
| 3 | 2 (3%) | 2 (10%) | 0 (0%) | 0.1148 | 1 (3%) | 1 (4%) | 0.9999 |
| History of CVA, n (%) | 10 (16%) | 3 (14%) | 7 (18%) | 0.7473 | 5 (16%) | 5 (19%) | 0.9999 |
| Direct Implant from ECMO, n (%) | 28 (46%) | 16 (76%) | 12 (30%) | ***0.0010** | 15 (47%) | 13 (48%) | 0.9999 |
| Disease Burden (in 1 week pre-implant) | | | | | | | |
| Mechanical Ventilation, n (%) | 48 (79%) | 17 (81%) | 31 (78%) | 0.9999 | 28 (88%) | 20 (74%) | 0.1871 |
| NMBA, n (%) | 26 (43%) | 7 (33%) | 19 (48%) | 0.4144 | 18 (56%) | 8 (30%) | ***0.0402** |
| TPN dependency, n (%) | 44 (72%) | 17 (81%) | 27 (68%) | 0.3709 | 23 (72%) | 21 (78%) | 0.7659 |
| Dialysis, n (%) | 5 (8%) | 1 (5%) | 4 (10%) | 0.6512 | 3 (9%) | 2 (7%) | 0.7869 |
| # of Inotropes, n (%) | | | | | | | |
| 1 | 23 (38%) | 7 (33%) | 16 (40%) | 0.7820 | 11 (34%) | 12 (44%) | 0.4295 |

| | | | | | | | |
|---|---|---|---|---|---|---|---|
| 2 | 30 (49%) | 10 (48%) | 20 (50%) | 0.9999 | 14 (44%) | 16 (59%) | 0.2352 |
| 3 | 4 (7%) | 1 (5%) | 3 (8%) | 0.9999 | 4 (13%) | 0 (0%) | 0.0571 |

Mann-Whitney U test for continuous variables and Chi-square test for categorical variables. *p<0.05.

** At the time of this study, one patient is alive on device, one has an unknown outcome. Both were excluded from analysis.

Wt: Weight, BSA: body surface area, RV: Right ventricle, LV: Left ventricle, INTERMACS: Interagency Registry for Mechanically Assisted Circulatory Support, CVA: cerebrovascular accident, ECMO: Extracorporeal membranous oxygenation, NMBA: neuromuscular blocking agent, TPN: total parenteral nutrition.
