## Supplementary material for "Oxygenator use in Ventricular Assist Devices from the Advanced Cardiac Therapies Improving Outcomes Network (ACTION) Registry": Table 2

**Table 2: On-Device Complications & Outcomes**

| Complications by # of occurrences (n of pts, %) Median [IQR] | Total pts n = 61 | In-Hospital Outcome | | P value | In-Hospital Outcome** | | P value |
|---|---|---|---|---|---|---|---|
| | | Early Oxygenator n = 21 | Late Oxygenator n = 40 | | Died n=32 | Transplanted or Recovered n=27 | |
| Device Days | 45 [16-163] | 113 [22.5-201] | 34.5 [12-87] | 0.1376 | 34 [16-87] | 97 [16-196] | 0.2234 |
| Total # of Adverse Events Reported | 157 (51, 84%) | 76 (17, 81%) | 81 (34, 85%) | 0.8373 | 101 (32, 100%) | 56 (19, 70%) | ***0.0001** |
| CNS events | 15 (13, 21%) | 9 (7, 33%) | 6 (6, 15%) | 0.0966 | 8 (7, 22%) | 7 (6, 22%) | 0.9744 |
| Ischemic CVA | 6 (6, 10%) | 4 (4, 14%) | 2 (2, 5%) | 0.0800 | 2 (2, 6%) | 4 (4, 15%) | 0.2782 |
| Hemorrhagic CVA | 2 (2, 3%) | 1 (1, 5%) | 1 (1, 3%) | 0.6374 | 1 (1, 3%) | 1 (1, 4%) | 0.9026 |
| Intracranial bleed | 7 (6, 8%) | 4 (3, 10%) | 3 (3, 8%) | 0.1778 | 6 (5, 16%) | 1 (1, 4%) | 0.1312 |
| Days, implant to CNS event | 14.5 [2.5-22.5] | 10 [2-20.5] | 22 [15-26.5] | 0.6121 | 11.5 [1-25] | 19 [6.5-22] | 0.5991 |
| Major Bleeding | 23 (22, 36%) | 7 (7, 33%) | 16 (15, 38%) | 0.7475 | 13 (14, 44%) | 10 (8, 30%) | 0.2638 |
| Post-op bleed | 6 (6, 10%) | 3 (3, 14%) | 3 (3, 8%) | 0.3978 | 0 (0, 0%) | 6 (6, 22%) | ***0.0034** |
| Cannulation Site | 1 (1, 2%) | 0 (0, 0%) | 1 (1, 3%) | 0.9999 | 1 (1, 3%) | 0 (0, 0%) | 0.9999 |
| Pulmonary | 5 (5, 8%) | 0 (0, 0%) | 5 (5, 13%) | 0.0908 | 5 (5, 16%) | 0 (0, 0%) | ***0.0318** |
| GI related | 8 (8, 13%) | 3 (3, 14%) | 5 (5, 13%) | 0.8444 | 4 (4, 13%) | 4 (4, 15%) | 0.9999 |
| Airway related | 3 (2, 3%) | 1 (1, 5%) | 2 (1, 3%) | 0.9999 | 2 (1, 3%) | 1 (1, 4%) | 0.9999 |
| Major Infection | 39 (22, 36%) | 23 (12, 57%) | 16 (10, 25%) | ***0.0130** | 29 (17 (53%) | 10 (5, 18%) | ***0.0062** |
| Blood stream, (n of pts, %) | 18 (13, 21%) | 10 (7, 33%) | 8 (6, 15%) | 0.0966 | 11 (8, 25%) | 7 (5, 18%) | 0.5496 |
| Pneumonia | 8 (6, 10%) | 4 (4, 19%) | 4 (2, 5%) | 0.0800 | 6 (4, 13%) | 2 (2, 7%) | 0.5191 |
| Mediastinitis, (n of pts, %) | 5 (5, 8%) | 2 (2, 10%) | 3 (3, 8%) | 0.7843 | 5 (5, 16%) | 0 (0, 0%) | ***0.0318** |
| Cannula site infection | 2 (2, 3%) | 1 (1, 5%) | 1 (1, 3%) | 0.9999 | 1 (1, 3%) | 1 (1, 4%) | 0.9999 |
| C. Difficile | 2 (1, 2%) | 2 (1, 5%) | 0 (0%) | 0.3443 | 2 (2, 6%) | 0 (0, 0%) | 0.4950 |
| NEC | 3 (2, 3% | 3 (2, 10%) | 0 (0, 0%) | 0.1148 | 3 (2, 6%) | 0 (0, 0%) | 0.4950 |
| Other localized infections | 3 (3, 5%) | 1 (1, 5%) | 2 (2, 5%) | 0.9999 | 2 (2, 6%) | 1 (1, 4%) | 0.5881 |
| Dialysis occurrences | 12 (12, 20%) | 6 (6, 29%) | 6 (6, 15%) | 0.2052 | 10 (10, 31%) | 2 (2, 7%) | ***0.0234** |
| Hepatic dysfunction | 5 (5, 8%) | 3 (3, 14%) | 2 (2, 5%) | 0.1761 | 4 (4, 13%) | 1 (1, 4%) | 0.2268 |
| Thrombus (non-CNS) | 3 (3, 5%) | 2 (2, 10%) | 1 (1, 3%) | 0.1999 | 1 (1, 3%) | 2 (2, 7%) | 0.5881 |
| Venous | 1 (2%) | 1 (1, 5%) | 0 (0, 0%) | - | 1 (1, 3%) | 0 (0, 0%) | - |
| Arterial | 2 (3%) | 1 (1, 5%) | 1 (1, 3%) | - | 0 (0, 0%) | 2 (2, 7%) | - |
| Hemolysis | 10 (9, 15%) | 8 (7, 33%) | 2 (2, 5%) | ***0.0057** | 7 (6, 19%) | 3 (3, 11%) | 0.4877 |
| Right heart failure | 4 (4, 7%) | 2 (2, 10%) | 2 (2, 5%) | 0.6024 | 1 (1, 3%) | 3 (3, 11%) | 0.3232 |
| Device malfunction | 8 (4, 7%) | 2 (2, 10%) | 6 (2, 5%) | 0.6024 | 2 (2, 6%) | 6 (2 7%) | 0.9999 |
| Hemorrhagic Pancreatitis | 1 (1, 2%) | 1 (1, 5%) | 0 (0, 0%) | 0.3443 | 1 (1, 3%) | 0 (0, 0%) | 0.9999 |
| Patient Outcome, n (%) | | | | | | | |
| Died on device | 32 (52%) | 11 (52%) | 21 (53Fmw%) | 0.9929 | - | - | - |
| Oxygenator removed | 28 (46%) | 10 (48%) | 18 (45%) | 0.8454 | - | - | - |
| Survived to Transplant or recovered | 27 (44%) | 10 (48%) | 17 (42%) | 0.7021 | - | - | - |
| Unknown outcome | 1 (2%) | 0 (0%) | 1 (3%) | - | | | |

Mann-Whitney U test for continuous variables, Fisher exact test for categorical variables, and logistic regression for event rate comparisons. *$p<0.05$. Table does not depict the 157 adverse events in their entirety. The most significant events are highlighted.

** At the time of this study, one patient is alive on device, one has an unknown outcome. Both were excluded from analysis.

ECMO: extracorporeal membrane oxygenation, GI: gastrointestinal, CNS; central nervous system, CVA: Cerebrovascular accident, NEC: Necrotizing enterocolitis.
